# Clinical practices underlie COVID-19 patient respiratory microbiome composition and its interactions with the host

**DOI:** 10.1101/2020.12.23.20248425

**Authors:** Verónica Lloréns-Rico, Ann C. Gregory, Johan Van Weyenbergh, Sander Jansen, Tina Van Buyten, Junbin Qian, Marcos Braz, Soraya Maria Menezes, Pierre Van Mol, Lore Vanderbeke, Christophe Dooms, Jan Gunst, Greet Hermans, Philippe Meersseman, CONTAGIOUS collaborators, Els Wauters, Johan Neyts, Diether Lambrechts, Joost Wauters, Jeroen Raes

**Affiliations:** Laboratory of Molecular Bacteriology, Department of Microbiology and Immunology, Rega Institute, KU Leuven, Belgium; Center for Microbiology, VIB, Leuven, Belgium; Laboratory for Clinical and Evolutionary Virology, Department of Microbiology and Immunology, Rega Institute, KU Leuven, Belgium; Laboratory of Virology and Chemotherapy, Department of Microbiology, Immunology and Transplantation, Rega Institute, KU Leuven, Belgium; Laboratory of Translational Genetics, Department of Human Genetics, KU Leuven, Belgium; VIB Center for Cancer Biology, VIB, Leuven, Belgium; Department of Pneumology, University Hospitals Leuven, Belgium; Laboratory of Clinical Bacteriology and Mycology, Department of Microbiology, Immunology and Transplantation, KU Leuven, Belgium; Laboratory of Respiratory Diseases and Thoracic Surgery (BREATHE), Department of Chronic Diseases and Metabolism, KU Leuven, Belgium; Laboratory of Intensive Care Medicine, Department of Cellular and Molecular Medicine, KU Leuven, Belgium; Laboratory for Clinical Infectious and Inflammatory Disorders, Department of Microbiology, Immunology and Transplantation, KU Leuven, Belgium

**Keywords:** COVID-19, SARS-CoV-2, respiratory microbiome, single-cell RNA-sequencing, host-microbiome interactions

## Abstract

Understanding the pathology of COVID-19 is a global research priority. Early evidence suggests that the respiratory microbiome may be playing a role in disease progression, yet current studies report contradictory results. Here, we examine potential confounders in COVID-19 respiratory microbiome studies by analyzing the upper (n=58) and lower (n=35) respiratory tract microbiome in well-phenotyped COVID-19 patients and controls combining microbiome sequencing, viral load determination, and immunoprofiling. We found that time in the intensive care unit and the type of oxygen support, both of which are associated to additional treatments such as antibiotic usage, explained the most variation within the upper respiratory tract microbiome, while SARS-CoV-2 viral load had a reduced impact. Specifically, mechanical ventilation was linked to altered community structure, lower species- and higher strain-level diversity, and significant shifts in oral taxa previously associated with COVID-19. Single-cell transcriptomic analysis of the lower respiratory tract of mechanically ventilated COVID-19 patients identified specific oral bacteria, different to those observed in controls. These oral taxa were found physically associated with proinflammatory immune cells, which showed higher levels of inflammatory markers. Overall, our findings suggest confounders are driving contradictory results in current COVID-19 microbiome studies and careful attention needs to be paid to ICU stay and type of oxygen support, as bacteria favored in these conditions may contribute to the inflammatory phenotypes observed in severe COVID-19 patients.

## Introduction

COVID-19, a novel coronavirus disease classified as a pandemic by the World Health Organization, has caused over 150 million reported cases and 3 million deaths worldwide to date. Infection by its causative agent, the novel coronavirus SARS-CoV-2, results in a wide range of clinical manifestations: it is estimated that around 80% of infected individuals are asymptomatic or present only mild respiratory and/or gastrointestinal symptoms, while the remaining 20% develop acute respiratory distress syndrome requiring hospitalization and oxygen support and, of those, 25% of cases necessitate critical care. Despite a concerted global research effort, many questions remain about the full spectrum of the disease severity. Independent studies from different countries, however, agree that age and sex are the major risk factors for disease severity and patient death^1–3^, as well as type 2 diabetes and obesity^4,5^. Other potential risk factors for critical condition and death are viral load of the patient upon hospital admission^6–8^ and the specific immune response to infection, with manifestation of an abnormal immune response in critical patients characterized by dysregulated levels of pro-inflammatory cytokines and chemokines, which some studies have associated with organ failure^9,10^.

Despite its close interplay with the immune system and its known associations with host health, little is known about the role of the respiratory microbiota in modulating COVID-19 disease severity, or its potential as a prognostic marker^11^. Previous studies exploring other pulmonary disorders have shown that lung microbiota members may exacerbate symptoms and contribute to their severity^12^, potentially through direct crosstalk with the immune system and/or due to bacteremia and secondary infections^13^. First studies of the respiratory microbiome in COVID-19 have revealed elevated levels of opportunistic pathogenic bacteria^14–16^. However, reports on bacterial diversity are contradictory. While some studies report a low microbial diversity in COVID-19 patients^14,17^ that rebounds following recovery^15^, others show an increased diversity in the COVID-19 associated microbiota^16^. These conflicting results could be due to differences in sampling location (upper or lower respiratory tract), severity of the patients, disease stage, treatment or other confounders. While these early findings already suggest that the lung microbiome could be exacerbating or mitigating COVID-19 progression, exact mechanisms are yet to be elucidated. Therefore, an urgent need exists for studies identifying and tackling confounders in order to discern true signals from noise.

To identify potential associations between COVID-19 severity and evolution and the upper and lower respiratory tract microbiota, we used nasopharyngeal swabs and bronchoalveolar lavage (BAL) samples, respectively. For the upper respiratory tract, we longitudinally profiled the nasopharyngeal microbiome of 58 COVID-19 patients during intensive care unit (ICU) treatment and after discharge to a classical hospital ward following clinical improvement, in conjunction with viral load determination and nCounter immune profiling. For the lower respiratory tract, we profiled microbial reads in cross-sectional single-cell RNA-seq data^18^ from of bronchoalveolar lavage (BAL) samples of 22 COVID-19 patients and 13 pneumonitis controls with negative COVID-19 qRT-PCR, obtained from the same hospital. The integration of these data enabled us to (1) identify potential confounders of COVID-19 microbiome associations, (2) explore how microbial diversity evolves throughout hospitalization, (3) study microbe-host cell interaction and (4) substantiate a link between the respiratory microbiome and SARS-CoV-2 viral load as well as COVID-19 disease severity. Altogether, our results suggest the existence of associations between the microbiota and specific immune cells in the context of COVID-19 disease. These interactions may be driven by mechanical ventilation and its associated clinical practices, and therefore could potentially influence COVID-19 disease progression and resolution.

## Results

### The upper respiratory microbiota of COVID-19 patients

We longitudinally profiled the upper respiratory microbiota of 58 patients diagnosed with COVID-19 based on a positive qRT-PCR test or a negative test with high clinical suspicion based on symptomatology and a chest CT-scan showing typical round glass opacities. All these patients were admitted and treated at UZ Leuven hospital. Patient demographics for this cohort are shown in Table 1.

**Table 1.**
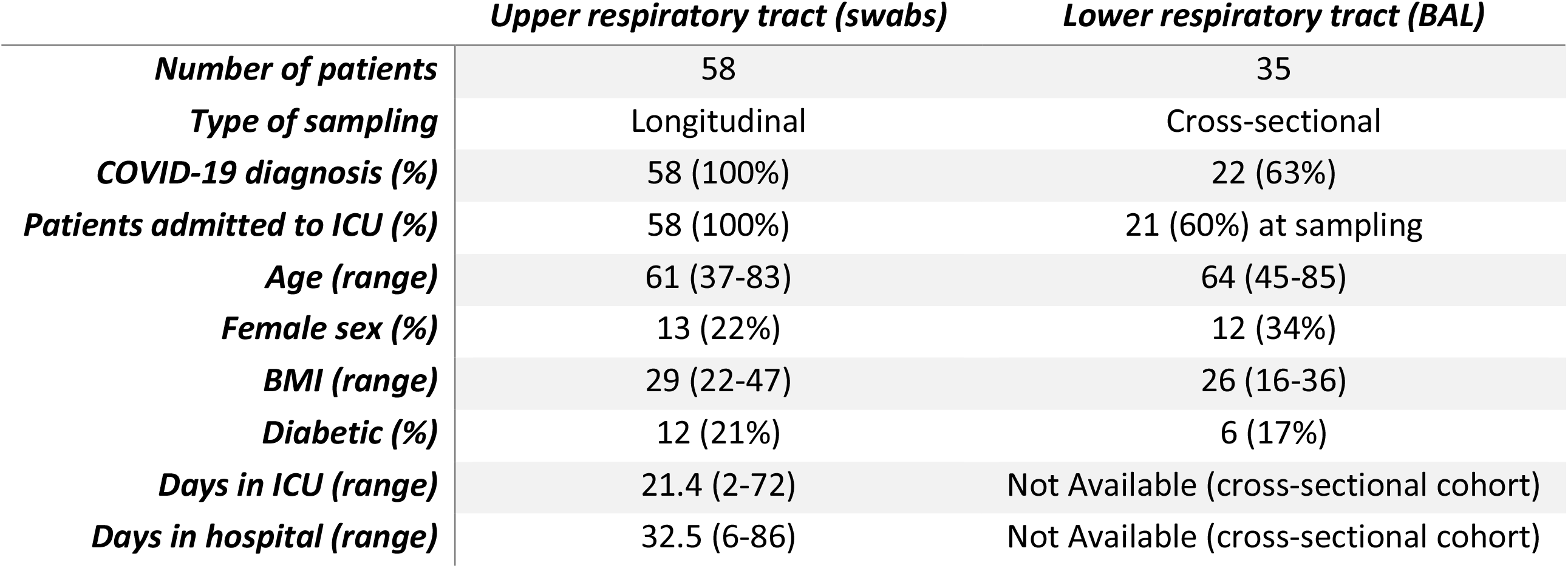
Patient demographics of upper and lower respiratory tract cohorts.

In total, 112 nasopharyngeal swabs from these patients were processed (Figure 1a): the V4 region of the 16S rRNA gene amplified on extracted DNA using 515F and 806R primers, and sequenced on an Illumina MiSeq platform (see Methods). From the same swabs, RNA was extracted to determine SARS-CoV-2 viral loads and to estimate immune cell populations of the host and expression of immune-related genes using nCounter (Methods). Of the 112 samples processed and sequenced, 101 yielded over 10,000 amplicon reads that could be assigned to bacteria at the genus level (Figure 1b; Methods). The microbiome of the entire cohort was dominated by the gram-positive genera *Staphylococcus* and *Corynebacterium*, typical from the nasal cavity and nasopharynx^19^.

**Figure 1.**
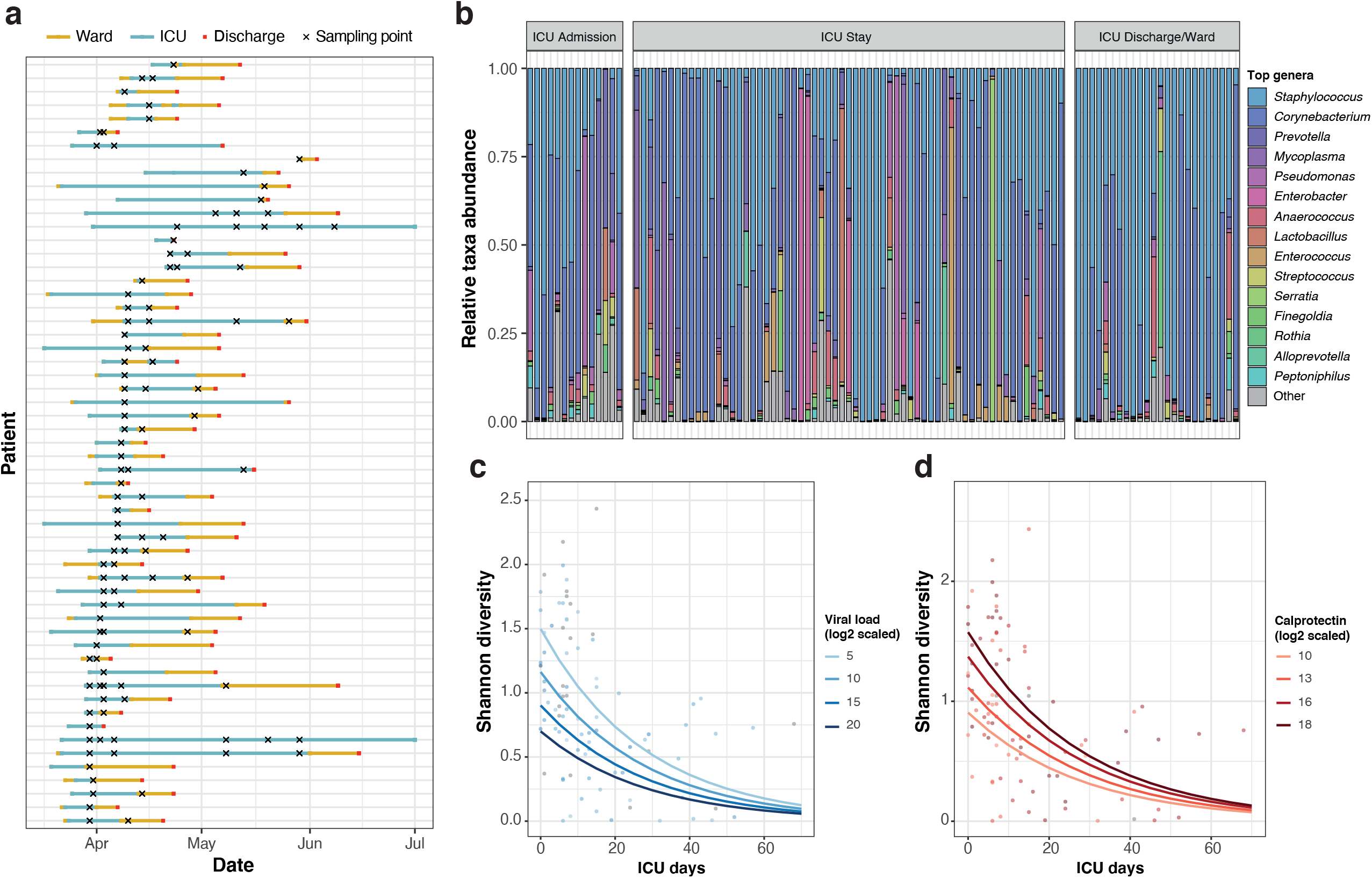
Sample overview and alpha diversity. a) Longitudinal sampling of patients. Each line represents one patient. Yellow lines span the days spent in ward, while blue lines span the days spent in ICU. Red points mark hospital discharge dates. Crosses indicate the timepoints where swab samples were obtained for microbiome analyses. b) Top 15 most abundant genera in this cohort. Samples with > 10,000 reads assigned to microbial taxa at the genus level were stratified according to the sampling moment: upon admission, throughout the ICU stay or at ICU discharge/during treatment in ward. c) Effect of the length of ICU stay and SARS-CoV-2 viral load on upper respiratory tract microbiome diversity. The plot shows the model-predicted Shannon index as a function of the days in ICU, for different levels of SARS-CoV-2 viral load (selected within the range of observed data). Confidence intervals for the predictions are shown in Supplementary Figure 1b. d) Association of the length of ICU stay and calprotectin gene expression levels with upper respiratory tract microbiome diversity. The plot shows the model-predicted Shannon index as a function of the days in ICU, for different levels of calprotectin (subunit S100A8) gene expression, selected within the range of observed data. Confidence intervals for the predictions are shown in Supplementary Figure 1c.

### Bacterial alpha diversity is associated with ICU stay length, SARS-CoV-2 viral load and calprotectin levels

First, we determined genus-level alpha-diversity for the 101 samples with more than 10,000 genus-level assigned reads, using the Shannon Diversity index (SDI; see Methods; Supplementary Table 1). We observed that the SDI was significantly different across sampling moments (Kruskal-Wallis test, p-value = 0.009; Supplementary Figure 1a), with significant differences between swabs procured upon patient ICU admission and later timepoints, suggesting an effect of disease progression and/or treatment (for instance due to antibiotics administered throughout ICU stay). We explored these differences further, and observed that SDI correlated negatively with the number of days spent in ICU at the moment of sampling, with longer ICU stays leading to a lower diversity (ρ=-0.53, p-value=1.9·10^−8^).

To evaluate the association of other clinical or disease-related variables with upper respiratory tract microbiome diversity, we used a generalized linear mixed model framework: we performed an exhaustive screening of all possible models containing up to 8 different explanatory variables, using an automated model selection algorithm (see Methods). The variables used to regress the SDI comprise the patient ID, modeled as a random effect; disease-related variables, such as the time in ICU, SARS-CoV-2 viral load or the use of mechanical ventilation; and other variables known to affect the microbiome, such as the administration of antibiotics (specifically meropenem/piperacillin-tazobactam and ceftriaxone) or the levels of inflammatory markers (calprotectin, C-reactive protein). The antibiotics meropenem and piperacillin/tazobactam were grouped as a single variable in all subsequent analyses as they were administered under the same clinical guidelines. The best performing model (AICc=121.79; p-value=4.06·10^−8^) included the patient modeled as a random effect and confirmed a negative association between the time spent in ICU and diversity. Additionally, this model showed a negative effect of SARS-CoV-2 viral load and a positive association of calprotectin levels with the SDI (Figure 1c,d; Supplementary Figure 1b-d).

We leveraged all the models generated in the screening to calculate weighted importance scores for all the fixed effects tested (Methods; Supplementary Figure 1e). These scores showed that the three variables incorporated in the best model (time in ICU, SARS-CoV-2 viral load and calprotectin) held the highest relative importance, followed by CRP levels and mechanical ventilation. Treatment with antibiotics ceftriaxone an meropenem or piperacillin-tazobactam had the lowest importance scores, and no significant differences in SDI were found between samples obtained before and after the administration of meropenem/piperacillin-tazobactam (Supplementary Figure 1f).

Altogether, our data suggest that respiratory microbiome diversity is linked to the length of ICU stay, SARS-CoV-2 viral load and calprotectin levels. While no significant effects were found for the most widely used antibiotics in this cohort, we cannot rule out that antibiotic administration or other clinical practices are causing the decrease of SDI over time.

### Respiratory microbiome composition variation is linked to respiratory support and associated clinical practices

We next explored potential associations between the upper respiratory genus-level microbiota composition and the extensive metadata collected in the study. In total, 72 covariates related to patient anthropometrics, medication and clinical variables measured in the hospital, as well as SARS-CoV-2 viral load, host cytokine expression and estimated immune cell populations measured in the swabs were tested (Supplementary Table 2). Individually, 20 of these covariates showed a significant correlation to microbiota composition in a univariate analysis (dbRDA, p-value<0.05; FDR<0.05; Figure 2a). These significant covariates were related to disease and measures of its severity, such as the clinical evaluation of the patient, the total length of the ICU stay, the number of days in ICU at the time of sampling, or the type of oxygen support needed by the patient. Despite showing an association to the overall diversity, SARS-CoV-2 viral load detected in the swabs was not significantly associated to microbiome composition variation (Supplementary Table 2). Neither ongoing antibiotic usage (i.e., administration of any type of antibiotic) nor number of ongoing antibiotics administered were significant, but the administration of specific antibiotics meropenem/piperacillin-tazobactam (previous or ongoing treatment) and ceftriaxone (ongoing administration only) showed significant associations with microbiome composition (Supplementary Table 2, Figure 2a).

**Figure 2.**
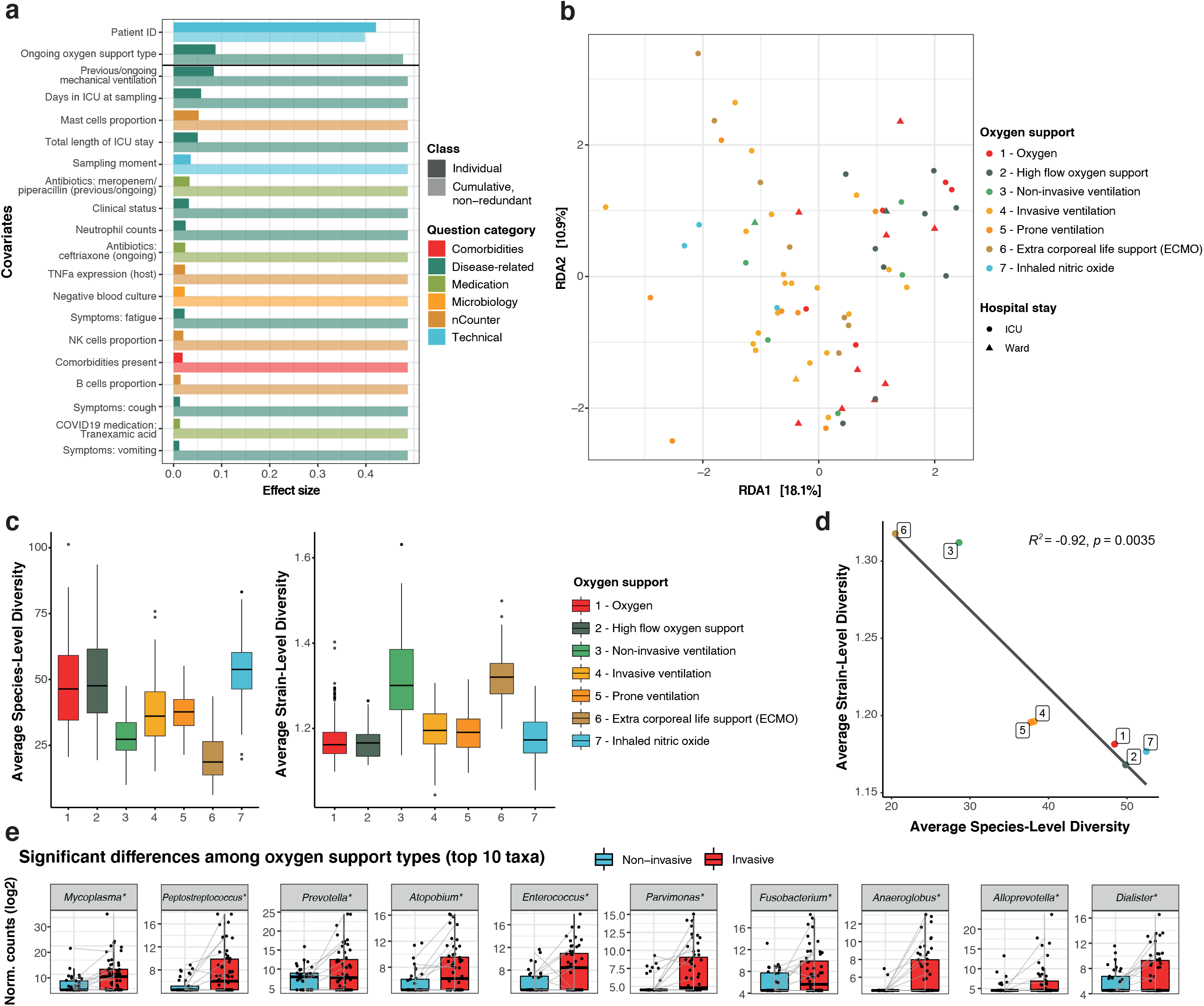
Upper respiratory microbiome covariates in COVID-19. a) Significant (BH-corrected p-value < 0.05) covariates explaining microbiota variation in the upper respiratory tract in this cohort. Individual covariates are listed on the y-axis, their color corresponds to the metadata category they belong to: technical data, disease-related, microbiological tests, comorbidities or host cell populations or gene expression, the latter measured with nCounter (see Methods). Darker colors refer to the individual variance explained by each of these covariates assuming independency, while lighter colors represent the cumulative and non-redundant variance explained by incorporating each variable to a model using a stepwise dbRDA analysis (using Aitchison distances). The black horizontal line separates those variables that are significant in the non-redundant analysis on top (Patient ID and oxygen support type) from the rest. b) RDA ordination plot showing the first 2 constrained axes. Ordination is constrained by the two significant variables “Patient ID” and “Oxygen support”. Samples are depicted as points, whose color indicates the oxygen support type of the patient and whose shape indicates stay at ward or ICU (at the moment of sampling). Axes indicate the variance explained by the first two constrained components of the RDA analysis. c) Species-(left) and strain-level diversity (right) of the samples, stratified by oxygen support type. d) Pearson correlation between average species- and strain-level diversity for each of the oxygen support categories. e) Significant differences in taxa abundances among oxygen support types. Differentially abundant taxa between invasive (red) and non-invasive (blue) ventilated samples. Only the top 10 most significant taxa are shown, as determined by their BH-adjusted p-value. Boxplots span from the first until the third quartile of the data distribution, and the horizontal line indicates the median value of the data. The whiskers extend from the quartiles until the last data point within 1.5 times the interquartile range, with outliers beyond. Individual data points are also represented. Gray lines join samples pertaining to the same patient, taken at different time points. Asterisks (*) indicate taxa that remain significant after controlling for the main antibiotics (ceftriaxone and meropenem/piperacillin-tazobactam).

Of the 20 significant covariates, only 2 accounted for 48.7% non-redundant variation in this dataset in a multivariate analysis (dbRDA; p-value=0.001), with the rest holding redundant information. These were the patient ID, included due to the longitudinal sampling of patients, and confirming that intra-individual variation over time is smaller than patient inter-individual variation^20^, and the type of oxygen support received at the time of sampling (Figure 2a,b). Notably, the type of oxygen support discriminated samples based on ventilation type, with non-invasive ventilation samples (groups 1, 2 and 3) separating from samples from intubated patients (groups 4 to 7; PERMANOVA test, R^2^=0.0642, p-value=0.001). Because of this separation, we also evaluated whether previous mechanical ventilation (regardless of the specific group) had a significant impact on the microbiome composition, showing even a larger effect size than when considering only the ongoing mechanical ventilation (PERMANOVA test, R^2^=0.0965, p-value=0.001), suggesting that this invasive procedure may have an effect that is prolonged in time.

Mechanical ventilation is inherently associated to additional clinical practices, such as administration of broad-spectrum antibiotics and decontamination procedures (including chlorhexidine washes) to prevent/treat ventilator-associated pneumonia. Hence, we explored whether antibiotic usage could explain the significant relationship between microbiome composition and oxygen support type. We found that from the specific antibiotics associated to microbiome composition, ceftriaxone was predominantly administered in patients on non-invasive oxygen support (Chi-square, p-value=0.001), whilst meropenem or piperacillin-tazobactam were preferentially given to patents on mechanical ventilation (Chi-square test, p-value=0.002; Supplementary Figure 2a). This association is not casual and responds to current treatment guidelines at UZ Leuven: ceftriaxone is administered to patients upon admission and for 3-7 days to prevent potential bacterial co-infections. In our cohort, 80% of the patients received ceftriaxone at the beginning of their stay (Supplementary Figure 2b). Patients with longer ICU stays and requiring higher levels of oxygen support will be considered to have hospital-acquired/ventilator-associated pneumonia (HAP/VAP) and receive meropenem or piperacillin/tazobactam (Supplementary Figure 2b). Therefore, the observed correlation between oxygen support types and these antibiotics can be explained by disease severity and length of ICU stay.

We therefore explored whether we could observe an effect of oxygen support type alone, deconfounding for the patient ID and the two significant antibiotic covariates using partial dbRDA to extract the effect size of oxygen support alone. The deconfounded model exhibited a significant association to overall microbiome composition (partial dbRDA; R^2^=0.058, p-value=0.042) suggesting that although antibiotic administration may explain part of the variation in microbiome composition observed, there may an independent effect of the oxygen support type. Nevertheless, the effect of other practices concomitant to mechanical ventilation, such as oral decontamination with chlorhexidine washes, could not be disentangled as these treatments were always performed together.

To determine if oxygen support or associated practices also impacted the microbiome at finer taxonomic resolution, we revisited alpha-diversity at species- and strain-level. We defined species as 97% identity 16S OTUs and strains per species as the clustered 16S sequences within each OTU. Our analyses revealed both species- and strain-level diversity change with ventilation, even with non-invasive ventilation (e.g. BIPAP, CPAP). Across all samples we observed high species- and low strain-level diversity pre-ventilation, which reversed following any form of ventilation (Figure 2c; Wilcoxon test; p-values<0.05, with the exception of type 7), with the exception of ventilation with inhaled nitric oxide. Further, species- and strain-level diversity showed a strong inverse correlation (Figure 2d; Pearson’s correlation, R^2^ = −0.92, p-value = 0.0035).

Given the observed effect of mechanical ventilation on the overall microbiome composition, we evaluated which specific taxa were differentially abundant between samples from intubated and non-intubated patients. In total, 28 genera were more abundant in samples from mechanically ventilated patients, while 1 genus was more abundant in non-invasively ventilated patients (p-value<0.05; FDR<0.05; Figure 2e, Supplementary Figure 3a; Supplementary Table 3). When controlling for the effect of the antibiotics ceftriaxone and meropenem/piperacillin, 20 genera were significantly different between both groups of samples (Supplementary Figure 3b, Supplementary Table 3). Some of these taxa are common oral microbiome commensals or opportunistic pathogens that had been repeatedly reported as more abundant in COVID-19 patients than in controls, such as *Prevotella, Fusobacterium, Porphyromonas* or *Lactobacillus*^14–16^. Here, we reported higher abundance of these genera in mechanically ventilated COVID-19 patients as compared to non-mechanically ventilated COVID-19 patients. This points at mechanical ventilation (and associated practices such as oral decontamination) as a potential confounder of previous COVID-19 studies. Additionally, we found other taxa not previously reported in previous COVID-19 microbiome studies, such as *Mycoplasma* or *Megasphaera* (Figure 2e, Supplementary Figure 2), but previously associated to risk of ventilator-associated pneumonia^21^.

By extracting the amplicon sequence variants (ASVs) corresponding to these differentially abundant genera (see Methods), some of these taxa could be narrowed down to the species level, confirming their origin as typically oral bacteria: for instance, *Prevotella* species included *P. oris, P. salivae, P. denticola, P. buccalis* and *P. oralis*. Within the *Mycoplasma* genus, ASVs were assigned to *Mycoplasma salivarium* among other species, an oral bacterium which has been previously associated to the incidence of ventilator-associated pneumonia^21^. When controlling for ventilation type, no taxa were found associated to SARS-CoV-2 viral loads (Supplementary Table 3). These results show that further research with larger cohorts and controlling for the relevant confounders highlighted here, such as ventilation type, antibiotic usage or length of stay in ICU, will be needed to study the specific effect of the viral infection.

### Single-cell RNA-seq of bronchoalveolar fluid identifies oral commensals and opportunist pathogens in the lower respiratory tract

Next, we explored what the functional consequences of (disease and/or treatment-driven) lung microbiome disturbances could be. To do so, we screened host single-cell RNA-seq data generated on BAL samples of 35 patients^18^ using a computational pipeline to identify microbial reads (see Methods). All patients in this cross-sectional cohort showed clinical symptoms of pneumonia, 22 of them being diagnosed with COVID-19. The other 13 patients with non-COVID-19 pneumonia were hereafter referred to as controls (Table 1). Out of the 35 patients, 21 were admitted to ICU (20 COVID-19 patients and 1 control) and 14 were hospitalized in ward at the moment of sampling (2 COVID-19 patients and 12 controls; Table 1 and Supplementary Figure 4). Microbial read screening of these samples revealed an average of 7,295.3 microbial reads per sample (ranging from 0 to 74,226 reads, with only a single sample yielding zero microbial reads; Supplementary Figure 4).

Among the top taxa encountered in these patients, we found similarities with the data obtained in nasopharyngeal swabs. The top 15 species detected include *Mycoplasma salivarium* as the dominating taxon in 5 COVID-19 patients in ICU, as well as different *Prevotella* members. Non-COVID-19 pneumonia patients in ward (i.e. non-mechanically ventilated) harbored different microbes: 2 patients had a microbiome dominated by *Porphyromonas gingivalis*, while a single patient had a microbiome dominated by the fungus *Pneumocystis jirovecii*, a known pathogen causing *Pneumocystis* pneumonia (PCP)^22^.

Supplementary table 4 shows associations between organism abundances and specific patient metadata: disease, hospital stay and ventilation type. Multiple links with COVID-19 diagnosis were identified (Wilcoxon test, (noncorrected) p-value<0.05; see Methods) but due to the low sample number, none was significant after multiple-test correction. Additionally, as hospital stay (ICU or ward), type of oxygen support (invasive or non-invasive ventilation) and disease (COVID-19 or controls) were highly correlated (Chi-squared test, p-value < 0.0001 for all three pairwise correlations), the effect of these three variables could not be disentangled. Therefore, although this data may validate our findings from the upper respiratory tract microbiome, due to the small cohort size and the existence of multiple confounders, these association results should be confirmed in larger studies.

### Bacteria in the lower respiratory tract associate to host cells from the innate immune system in COVID-19 patients

Next, we took advantage of the single-cell barcoding and questioned whether the microbial reads that we identified were found in association with host cells (for instance infecting or internalized), or contrarily, had unique barcodes suggesting a free-living state. In total, 29,886 unique barcodes were identified that matched a total of 46,151 microbial UMIs. The distribution of UMIs per barcode was asymmetrical, ranging from 1 to 201 and with 88% of the barcodes having a single UMI. Additionally, 26,572 barcodes (89%) were associated to a single microbial species, the rest being associated to 2 species (8.8%) or more (2.2%).

Out of the total 29,886 microbial barcodes, only 2,108 were also assigned to host cells, suggesting that the bulk of bacteria found in BAL samples exist as free-living organisms or in bacterial biofilms. Although microscopic evaluation would be needed to validate this hypothesis, bacterial biofilms have been previously documented in bronchoalveolar lavages^23^, and the enrichment for host cells in these samples via centrifugation^18^ may have also indirectly enriched these specimens for biofilm and/or host-associated microbes. However, for the fraction of bacteria associated to host cells, the distribution across disease types was not random. We found that while 2.3% of the non-COVID-19 patient cells were associated to bacterial cells, almost the double (4%) could be observed in COVID-19 patients (Figure 3a; Chi-squared test; p-value < 2.2·10^−16^). However, because COVID-19 diagnosis is highly correlated with mechanical ventilation in this cohort, this effect could be due to higher intubation rates in COVID-19 patients and possibly, a higher incidence of VAP. Within COVID-19 patients, we also evaluated the overlap between bacteria-associated host cells and cells with detected SARS-CoV-2 reads^18^ (Supplementary Table 5). Out of 1,033 host cells associated with bacteria in COVID-19 patients and 343 cells with detected SARS-CoV-2 reads, only one cell was positive for both viral and bacterial reads. A binomial test for independence of virus and bacteria detection in the same host cell, showed that the observed co-occurrence in one cell only was highly unlikely (p-value=5.7·10^−4^), therefore suggesting mutual exclusion of microbiome members and viruses in the same host immune cells. However, it must be noted that lack of detection does not necessarily imply lack of association of bacteria/virus to host cells, especially with experimental methods such as single-cell RNA-seq, designed for profiling host cells and not optimized for detection of these entities. Therefore, further studies with larger sample sizes are required to validate the co-exclusion hypothesis.

**Figure 3.**
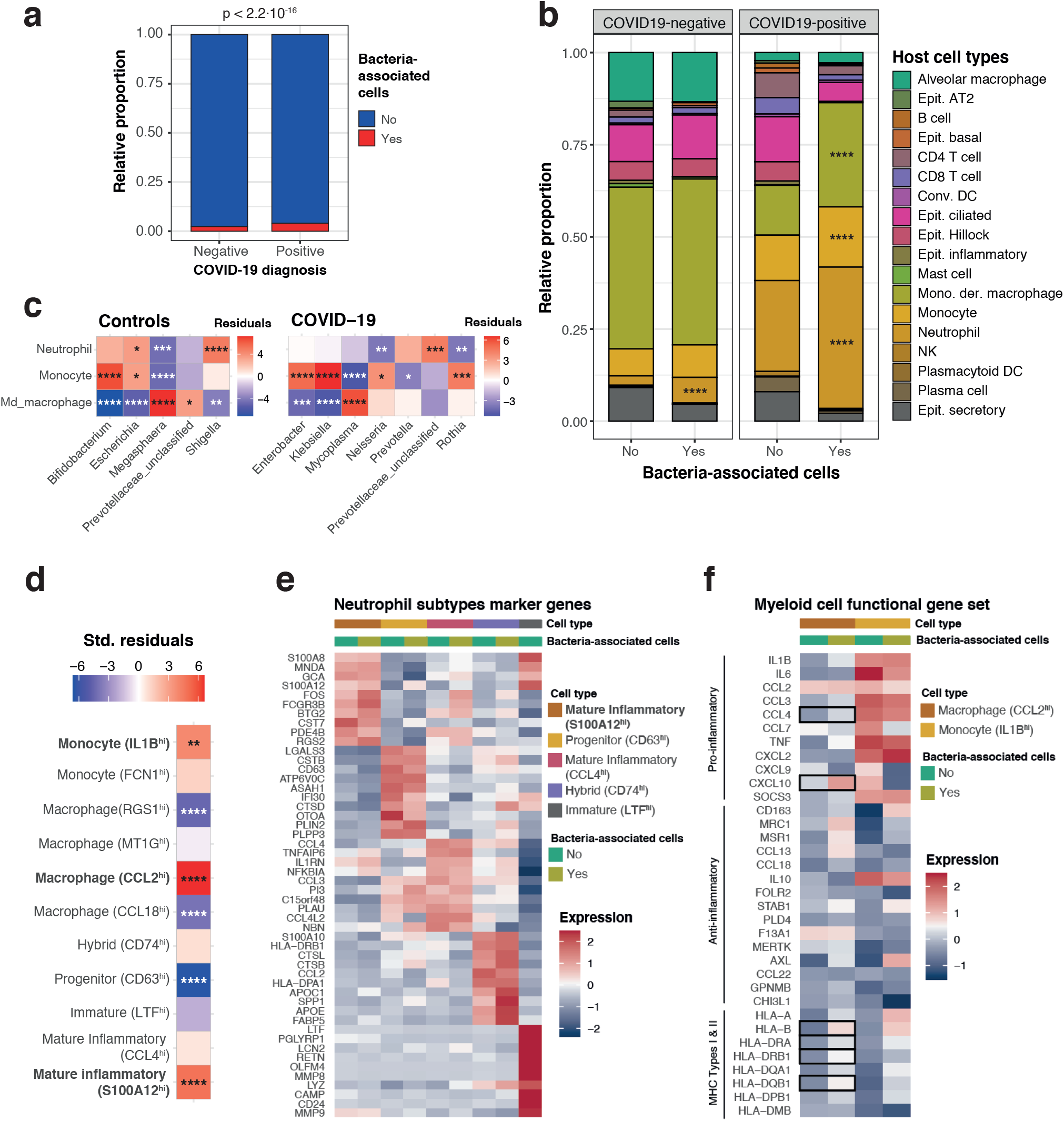
Host single cells associated to the lower respiratory tract microbiota. a) relative proportion of cells from negative and positive COVID-19 patients with (red color) and without (blue) associated bacteria. The p-value of a chi-squared test using the count data is shown on top of the panel. b) Cell types enriched in bacteria-associated cells. Barplots represent the proportion of cell types without (“No”) and with (“Yes”) bacteria in COVID-19 positive and negative patients. For each patient class, we tested for enrichment of bacteria-associated cells (“Yes”) across the different cell types, using the proportions of non-bacteria associated cells (“No”) as background. Asterisks mark the cell types with significant enrichment of bacteria. c) Bacterial genera preferentially associating to specific cell types. The heatmaps show the standardized residuals of a chi-squared test including all bacterial genera and the three host cell types enriched in bacteria, for controls (left) and COVID-19 positive patients (right). Taxa with no significant associations with any of the cell types are not shown. Asterisks denote significant positive or negative associations: enrichments are shown in red; depletions are depicted in blue. d) Host cell subtypes associated with bacteria. The heatmap shows the standardized residuals of a chi-squared test including the subtypes of neutrophils, monocytes and monocyte-derived macrophages with associated bacteria, considering cells without bacteria as background. Asterisks denote significant positive or negative associations: enrichments are shown in red; depletions are depicted in blue. e) Marker genes detected for the 5 different subtypes of neutrophils. The heatmap also shows within-group differences between bacteria-associated and bacteria-non-associated cells. f) Myeloid cell functional gene set showing the expression of canonical pro-inflammatory, anti-inflammatory and MHC genes for the two subtypes of myeloid cells significantly associated with bacteria (CCL2^hi^-macrophages and IL1B^hi^-monocytes). The heatmap also shows within group differences between bacteria-associated and bacteria-non-associated cells. Statistically significant differences after multiple testing correction are marked with squares. For b)-d) asterisks denote significance as follows: * = p-value ≤ 0.05; ** = p-value ≤ 0.01; *** = p-value ≤ 0.001; **** = p-value ≤ 0.0001.

We also explored whether host-associated bacterial reads would preferentially be linked with specific cell types, taking into account the varying frequencies of cell types in COVID-19 patients and controls (see Methods). Such a preferential association would suggest that these observations are biologically relevant and not an artifact of the single-cell sample and library preparation. Among control patients, cell types were similarly distributed in both groups (i.e. with and without bacteria), with only a preferential association of microbial cells with neutrophils (p-value = 3.61·10^−12^; Figure 3b; Supplementary Figure 4). However, in COVID-19 patients, three cell types were significantly associated with bacteria: neutrophils (p-value < 2.2·10^−16^), monocytes ((p-value = 4.82·10^−5^) and monocyte-derived macrophages (p-value < 2.2·10^−16^; Figure 3b; Supplementary Figure 5). We also found that different bacteria associate with distinct host cells. For instance, in COVID-19 patients, bacteria from the *Mycoplasma* genus preferentially associated to monocyte-derived macrophages (p-value = 2.28·10^−7^), while *Rothia* (p-value = 8.21·10^−4^), *Enterobacter* (p-value = 2.59·10^−5^), or *Klebsiella* (p-value = 3.12·10^−9^) are enriched in monocytes (Figure 3c).

Last, we investigated whether the associations of bacteria to host cells are linked to host cell expression. To do so, we assessed whether expression based cell subtype classification^18^ for neutrophils, monocytes and macrophages showed non-random associations with bacteria across all samples in this cohort. Among the neutrophils, a subtype of inflammatory neutrophils characterized by expression of the calgranulin S100A12 was enriched in bacteria-associated cells (p-value 7.18·10^−6^; Figure 3d,e). This subset of cells was also found to be enriched in SARS-CoV-2 nucleocapsid gene reads^18^, suggesting that the same cell type responsible for defense against the virus would be responding to potentially invasive bacteria in the lung. This subgroup is characterized by the expression of the calprotectin subunits S100A8 and S100A9. It is known that S100A8/A9 heterodimer secretion is increased in infection-induced inflammation and has some antibacterial effects mediated by secretion of pro-inflammatory cytokines, release of reactive oxygen species and recruitment of other inflammatory cells, as well as chelation of Zn^2+^ necessary for bacterial enzymatic activity^24^. These mechanisms are mediated by binding of the S100A8/A9 dimer to TLR4 receptors to trigger the release of pro-inflammatory cytokines such as IL-6 and TNF-α, and thus may contribute to sustain or exacerbate inflammation^25^. Therefore, the association with bacteria could, at least in part, explain the inflammatory phenotype of this neutrophil subset. To further examine this hypothesis, we explored differential gene expression between bacteria-associated and non-associated S100A12^hi^ neutrophils (Supplementary Table 6). Because association of these cells with SARS-CoV-2 and with bacteria was mutually exclusive, we also compared these changes with the ones triggered by the virus in neutrophils^26^. Within this subset, neutrophils with co-occurring bacteria showed significantly higher expression (Bonferroni-corrected p-value < 0.05) of pro-inflammatory genes, including the cytokine IL1B and some of its target genes (PTSG2), the transcription factors FOS and JUN, and several genes involved in degranulation (S100A9, FOLR3, HSPA1A, HSP90AA1, FCGR3B), (Supplementary Table 6). Among these, FOLR3, a gene encoding for a folate receptor, is found in neutrophil secretory granules and has antibacterial functions, by binding folates and thus depriving bacteria of these essential metabolites^27^. This response differed to that of virus-engulfing neutrophils in that IFN response genes are not distinctively upregulated by bacteria.

Regarding myeloid cells, both inflammatory IL1B^hi^ monocytes (p-value = 2·10^−16^) as well as a mixed group of CCL2-expressing macrophages (p-value = 5.38·10^−10^) are enriched in bacteria-associated cells (Figure 3f). These inflammatory monocytes are believed to have an important role in the aberrant immune response occurring in severe COVID-19 patients. In this case, further gene expression patterns were detected, specific for bacteria-associated cells: for CCL2^hi^ macrophages, cells with co-occurring bacteria expressed higher levels of MHC genes of type I and II, suggesting a more active role of these cells in antigen presentation (Bonferroni-corrected p-value < 0.05; Figure 3f; Supplementary Table 6). A similar increase was also observed in monocytes, yet not significant (Supplementary Table 6), possibly due to the lower monocyte abundances in this dataset. Additionally, bacteria-associated macrophages express significantly higher levels of the calprotectin subunits S100A8/A9, similarly to neutrophils, as well as pro-inflammatory chemokines (such as CCL4, CXCL10 and CXCL1).

Altogether, our results suggest that the bacteria detected in these cell subsets via scRNA-seq analyses may be contributing to the inflammatory response observed in the host.

## Discussion

Since the beginning of the COVID-19 pandemic, a massive global effort by the scientific community was undertaken to understand physiopathology of SARS-CoV-2 infection and risk factors affecting disease outcome. In this study, we explored the respiratory microbiota as a potential risk factor for disease severity, and we evaluated the upper and lower respiratory tract microbiota in COVID-19 patients throughout hospitalization. We linked this data to viral load measurements and immunoprofiling results from nCounter and single-cell RNA sequencing data. To assess robustness of previously reported signals, we investigated the effect of potential confounders based on a broad panel of patient metadata variables.

We found that in the upper respiratory tract, SARS-CoV-2 viral load has a mild negative association with bacterial biodiversity. A larger effect of severity indicators such as calprotectin levels or length of ICU stay was observed, with diversity decreasing throughout the length of the ICU period, a pattern reminiscent of that seen in other pulmonary conditions^28,29^. The effect of ICU length-of-stay may be mediated by treatment options such as the administration of broad-spectrum antibiotics and/or patient intubation and mechanical ventilation. Antibiotic usage might also explain why calprotectin levels correlate with alpha diversity: such antibiotics would decrease overall microbial diversity, including (some of) the taxa that could be linked to inflammation. The observed effects of these clinical practices on microbiome alpha diversity could potentially explain why previous studies on the microbiota of COVID-19 patients show conflicting results regarding diversity: some studies reported lower diversity in sputum or throat swab samples of COVID-19 patients^14,15,17^ while others focusing on the lower respiratory microbiome using bronchoalveolar fluid samples, showed higher bacterial diversity in COVID-19 patients than in controls^16^. To further complicate matters, it cannot be excluded that sampling site or processing could also be potential confounders in these studies and/or reflect the different pathologies in the different areas of the respiratory tract.

We further found that between-patient microbiome variation (as measured by genus-level microbial beta-diversity) was also influenced by different severity indicators such as the clinical status of the patient, or more importantly the type of oxygen support received, with mechanically ventilated patients harboring a different microbiota than non-intubated patients. This effect could not be fully explained by neither general antibiotic administration, nor the usage of specific antibiotics such as ceftriaxone, meropenem or piperacillin-tazobactam, suggesting an independent effect of mechanical ventilation. Such an independent effect has previously been suggested in small cohorts^28,30,31^, but it needs to be validated in larger studies. However, other associated practices such as decontamination procedures could still be responsible for the observed associations. The impact of oxygen support was also reflected at the species- and strain-levels, with intubation causing a significant decrease and increase, respectively, in diversity. We hypothesize that the introduction of forced oxygen may drive the fast extinction of certain microbial species enabling the diversification of existing or newly colonizing species into new strains. Combined, these results suggest that non-invasive ventilation (e.g. BIPAP, CPAP) can have microbial effects indicating that any form of ventilation may be a tipping point for microbial community differences.

Importantly, several of the taxa reported to change between intubated and non-intubated patients were reported to be linked to diagnosis in previous COVID-19 microbiome studies^14–16^. In our study, no taxa were specifically linked to SARS-CoV-2 viral load after controlling for mechanical ventilation. This result suggests the possibility that mechanical ventilation and its associated clinical practices are confounding previous results. Indeed, one study comparing COVID-19 patients with patients diagnosed of community-acquired pneumonia found no differences in respiratory microbiome composition between both groups of patients, but both groups did differ from healthy controls^32^. Together, these results indicate that patient intubation or even non-invasive ventilation, as well as their associated medical practices, are to be considered as important confounders when studying the upper respiratory microbiome, and we strongly suggest future COVID-19 microbiome studies should foresee and include strategies to account for this covariate. As an example, a recent study found a single ASV corresponding to the genus *Rothia* that was specific for SARS-CoV-2 patients after controlling for ICU-related confounders by comparing with a previous study of the microbiome in ICU patients^33^. Additionally, these findings on potential drivers of microbiome variation are not exclusive to COVID-19 disease: the effect of intubation on the respiratory microbiome and its influence on the incidence of ventilator-associated pneumonia have been previously studied^28,30,31^.

To better understand the potential functional consequences of these procedures and linked microbial shifts, we also profiled the microbiome of the lower respiratory tract using single-cell data obtained from a cross-sectional cohort of patients derived from the same hospital. Our results show that single-cell RNA-seq, despite not being optimized for microbial detection and profiling, can identify bacteria alone or in association with specific human cells. Unfortunately, the low numbers of microbial reads obtained in this small cohort, together with the fact that ICU stay, COVID-19 diagnosis and intubation are highly correlated in this set of patients, only allow for a first exploratory analysis of the results, requiring validation in further datasets. In this cohort, we identified different oral commensals and opportunistic pathogens previously linked to COVID-19 patients in both groups of samples, thus pointing again at a potential ventilation-linked origin. More interestingly, we identified a subset of bacteria associated with host cells, more specifically with neutrophils, monocytes and macrophages. This enrichment shows that these bacteria are likely not random contaminants, from which an even distribution across cell types (i.e. considering cell type abundances) would be expected. The identity of these host cells suggests that bacteria could have been phagocyted by these innate immune system cells, rather than be attached to the host cell surface. To the best of our knowledge, this is the first study linking interacting host cells and lung microbiome via high-throughput single-cell RNA-seq.

We find that host cells associated with bacteria, most of which are of oral origin, exhibit pro-inflammatory phenotypes as well as higher levels of MHC for antigen presentation. In this single-cell cohort it was observed that critical COVID-19 patients are characterized by an impaired monocyte to macrophage differentiation, resulting in an excess of pro-inflammatory monocytes, as well as by prolonged neutrophil inflammation^26^. Given that only these cell types are enriched in bacteria, we hypothesize that the respiratory (or ventilation-linked) microbiome may be playing a role in exacerbating COVID-19 progression in the lower respiratory tract. We verified that this response could likely be driven by bacteria and not SARS-CoV-2, which is also detected mostly in these cell types, as there is almost no overlap in detection of both virus and bacteria in the same cells. However, it must be noted that lack of detection does not completely rule out presence of virus and bacteria within these cells. Therefore, further research is required in order to confirm a causative role of the microbiota in this immune impairment characteristic of critical disease, and to reveal the specific mechanisms involved.

The presence of oral taxa in the lung microbiota is not unique of disease conditions. It is known that microaspiration, or the aspiration of aerosol droplets originated in the oral cavity, occurs in healthy individuals and can serve as a route for lung colonization of oral commensals^34^. Such an increase of oral bacteria in the lower respiratory tract could be facilitated when critically ill patients –including but not limited to COVID-19– get intubated. Indeed, oral bacteria have been linked to ventilator-associated pneumonia^35,36^. It is yet to be elucidated whether COVID-19 physiopathology favors lung colonization by oral bacteria or if, in contrast, a lung microbiome previously colonized by oral microbes could also contribute to the disease. What is known is that an increase of oral bacteria in the lower respiratory tract can result in an increased inflammatory phenotype, even in healthy subjects^37^

## Conclusion

Overall, this study provides a systematic analysis of potential confounders in COVID-19 microbiome studies. We identified that ICU hospitalization and type of oxygen support, which may be at least partially explained by clinical practices such as antibiotic usage, had large impacts on the upper respiratory tract microbiome and have the potential to confound clinical microbiome studies. Among the different types of oxygen support we reported the largest shifts in microbial community structure between intubated and non-intubated patients. We found that oral taxa were strongly enriched in the upper respiratory tract of mechanically ventilated COVID-19 patients, and specific taxa were also found in the lower respiratory tract of COVID-19 patients. Further, in the lower respiratory tract, microbes were strongly associated with specific pro-inflammatory immune cells. This information contributes to a collective body of literature on the pathology of COVID-19 and suggests that careful attention be paid to ICU stay and type of oxygen support and associated clinical practices such as antibiotic usage or oral decontamination procedures when evaluating the role of the lung microbiome on COVID-19 disease progression.

## Methods

### Study design and patient cohorts

All experimental protocols and data analyses were approved by the Ethics Commission from the UZ Leuven Hospital, under the COntAGIouS observational clinical trial (study number S63381). The study design is compliant with all relevant ethical regulations, including the Declaration of Helsinki and in the GDPR. All participants gave their informed consent to participate in the study.

A total of 58 patients from the COntAGIouS observational trial were included as our upper respiratory tract cohort. All patients were admitted to the UZ Leuven hospital with a diagnostic of COVID-19. The disease was diagnosed based on a) a positive qRT-PCR test, performed on admission or previously on other hospitals, when patients were transferred from other medical facilities; or b) a chest CT-scan showing alveolar damage and clinical symptoms of the disease. All patients included in the study were admitted to ICU for a variable amount of time. Nasopharyngeal swabs were taken from these patients at different timepoints throughout ICU stay and after ICU discharge, during recovery in ward. A total of 112 swabs were processed for upper respiratory microbiome characterization (Figure 1a).

To extend our findings from the upper respiratory tract, we also profiled the lower respiratory tract microbiota in a different cohort^18^ of 35 patients belonging to the same observational trial and also recruited at UZ Leuven hospital. This cross-sectional cohort is composed by 22 COVID-19 patients and 13 pneumonitis controls with negative qRT-PCR for SARS-CoV-2, with varying disease severity. Previous data from single-cell RNA-sequencing had been collected for this cohort^18^. We reanalyzed this single-cell dataset to profile the lower respiratory tract microbiota in these patients.

### RNA/DNA extraction and sequencing

Nucleic acid extraction from the swab samples was performed with AllPrep DNA/RNA/miRNA Universal kit (QIAGEN, catnr. 80224). Briefly, swabs from the potentially infectious samples were inactivated by adding 600µL RLT-plus lysis buffer. To increase bacterial cell lysis efficiency, glass beads and DX reagent (Pathogen Lysis Tubes, QIAGEN, catnr. 19091) were added to the lysis buffer, and samples were disrupted in a FastPrep-24™ instrument with the following program: 1-minute beating at 6.5m/sec, 1-minute incubation at 4°C, 1-minute beating at 6.5m/sec, 1-minute incubation at 4°C. After lysis, the remaining extraction steps followed the recommended protocol from the manufacturer. DNA was eluted in 50µL EB buffer. Amplification of the V4 region of the 16S gene was done with primers 515F and 806R, using single multiplex identifiers and adaptors as previously described^38^. RNA was eluted in 30µL of nuclease-free water and used for SARS-CoV-2 viral load determination in the swabs as well as to measure inflammatory markers and cytokines and to estimate host cell populations via marker gene expression using nCounter. In brief, raw nCounter data were processed using nSolver 4.0 software (Nanostring), sequentially correcting three factors for each individual sample: technical variation between samples (using spiked positive control RNA), background correction (using spiked negative control RNA) and RNA content variation (using 15 housekeeping genes). We have previously validated nCounter digital transcriptomics for simultaneous quantification of host immune and viral transcripts^39^, including respiratory viruses in nasopharyngeal aspirates, even with low RNA yield^40–42^.

DNA sequencing was performed on an Illumina MiSeq instrument, generating paired-end reads of 250 base pairs.

For quality control, reads were demultiplexed with LotuS v1.565^43^ and processed following the DADA2 microbiome pipeline using the R packages DADA2^44^ and phyloseq^45^. Briefly, reads were filtered and trimmed using the parameters truncQ=11, truncLen=c(130,200), and trimLeft=c(30, 30) and then denoised. After removing chimeras, amplicon sequence variants (ASVs) table was constructed and taxonomy was assigned using the Ribosomal Database Project (RDP) classifier implemented in DADA2 (RDP trainset 16/release 11.5). The abundance table was then corrected for copy number, rarefied to even sequencing depth, and decontaminated. For decontamination, we used the prevalence-based contaminant identification method in the R package decontam^46^.

### 16S statistical analysis

All the 16S data analyses were performed using R v3.6.0 and the packages vegan (v2.5.7)^47^, phyloseq (v1.34.0)^45^, CoDaSeq (v0.99.6)^48^, DESeq2 (v1.30.1)^49^, Biostrings (v2.58.0)^50^, rstatix (v0.7.0)^51^, glmulti (v1.0.8)^52^, sjPlot (v2.8.7)^53^, and DECIPHER (v2.18.1)^54^.

To analyze the 16S amplicon data, technical replicates were pooled and counts from technical replicates were added. For all the analyses using genus-level agglomerated data, only samples containing more than 10,000 reads assigned at the genus level were used (101 samples in total). Alpha-diversity was analyzed using Shannon’s Diversity Index. Comparison of the alpha diversity values across different groups was performed using Kruskal-Wallis tests for comparisons across multiple groups. When applicable, pairwise comparisons were performed using Dunn post-hoc tests. To establish the potential associations of alpha diversity with different metadata variables, we selected 8 variables related to COVID-19 disease and/or known to affect microbiome composition and diversity: patient ID, days spent in ICU, SARS-CoV-2 viral load, antibiotic usage for ceftriaxone and meropenem/piperacillin-tazobactam, previous mechanical ventilation, calprotectin gene expression and CRP levels. Meropenem and piperacillin-tazobactam were merged as a single antibiotic as their administration is indicated under the same clinical guidelines.

We used the R package glmulti to perform an exhaustive evaluation of the 256 models including all possible combinations of the selected variables. All models generated were generalized linear models or generalized linear mixed models (when including the patient ID as a random effect), using a Gaussian family with a logarithmic link. Model ranking and selection was performed based on the lowest small-sample-corrected Akaike Information Criterion (AICc), and model significance was assessed comparing with a null model (including the intercept only) using ANOVA test. Final variable importance was calculated as a weighted average of the models in which each of the variables appeared, with weights corresponding to the model ranks, defined by their AICc values. This was also implemented as part of the glmulti package. The final model plots were generated with the sjPlot package. Intra-patient differences in alpha diversity between timepoints before and after administration of antibiotics or mechanical ventilation were determined with Wilcoxon signed-rank tests.

Beta diversity analyses were performed using distance-based redundancy analyses (dbRDA), using Aitchison distances. Prior to CLR data transformation, we filtered the data using the CoDaSeq.filter function, to keep samples with more than 10,000 reads and taxa with a relative abundance above 0.1% in any sample, as well as a prevalence of at least 10% in the cohort. To replace zeros, we first calculated the minimum (non-zero) relative abundance of each taxon across all samples. Then, for samples with zero counts for a given taxon, the minimum relative abundance of the specific taxon was multiplied by the total counts of such samples and this value was used to impute the zeros. dbRDA analyses were performed using the capscale function from vegan, first in univariate analyses with 72 metadata variables (Supplementary Table 2). Model p-values were corrected using Benjamini-Hochberg’s (BH) multiple-testing correction, to select 20 variables with BH-adjusted p-values < 0.05. These 20 variables were included in a multivariate model, and non-redundant contribution to variation was calculated using forward stepwise variable selection via the ordiR2step function from vegan. To deconfound the effect of antibiotics and patient ID for oxygen support type, partial dbRDA was used, including both antibiotics and patient ID as blocking variables. Metadata variables containing dates, as well as non-informative metadata were excluded. Non-informative metadata variables were defined as those containing a single non-NA value or, for categorical variables, those being unevenly distributed (with >90% of the samples belonging to the same category, for instance an antibiotic administered only in two different samples). Additionally, from pairs of highly collinear variables (correlation higher than 0.9), only one variable was kept.

Differential taxa abundance analyses were performed using DESeq2’s likelihood ratio tests and controlling for potential confounders when indicated, including them in the null model.

To explore species-level and strain-level diversity, 16S sequences were first clustered into 97% nucleotide diversity operational taxonomic units (OTUs) using the R packages Biostrings and DECIPHER. These OTUs were used to represents the species-level. The number of unique 16S sequences clustered within each OTU were used to represent the number of detectable strains per species. To calculate strain-level diversity per sample, the number of strains of 5 detected OTU species were randomly selected and averaged. This was repeated 1,000x and the average of the all 1,000 subsamplings was used as the final strain-level diversity value for each sample, as previously described^55^. To account for uneven sampling assessing diversity differences based on different parameters, we randomly selected and averaged the species- and strain-level diversity of 5 samples per parameter. This was repeated 100x and the subsamplings were used to assess the significant differences between species- and strain-level diversity across the parameters. The average was of all 100 subsamplings was used to as the input for a Pearson’s correlation between species- and strain-level diversity.

All statistical tests were two-sided unless otherwise specified, and when multiple tests were applied to the different features (e.g. the differential taxa abundances across two conditions) p-values were corrected for multiple testing using Benjamini-Hochberg’s method.

### Identification of microbial reads in BAL scRNA-seq data

BAL scRNA-seq raw fastq data, as well as cell type and subtype assignations for all individual cells, were obtained from a previous publication from within the COntAGIouS consortium. Experimental procedures on BAL samples as well as detailed host single-cell gene expression analyses are detailed in the original publication^18^.

5’ single-cell RNA-seq data obtained from the 10X Genomics Chromium platform was processed with an in-house pipeline to identify microbial reads. This pipeline comprises a series of steps designed to detect bacterial reads with high sensitivity, while discarding potential false positives. For microbial identification, only the read 2 fastq file from the raw sequencing files, containing the information on the cDNA fragment, was used. Trimmomatic^56^ (v0.38) was used to trim low quality bases and adapters, and discard short reads. Additionally, Prinseq++^57^ (v1.2) was used to discard reads with low-complexity stretches such as poly-A sequences. Following these two quality control steps, reads from human and potential sequencing artifacts (phage phiX174) were mapped with STAR^58^ (v2.7.1) and discarded. The remaining unmapped reads were mapped against reference microbial genomes using a 2-step approach: first, we scanned these remaining reads using mash screen^59^ (v2.0) against a custom database of 11685 microbial reference genomes including bacteria, archaea, fungi and viruses. Genomes likely to be present in the analyzed sample (selected using a threshold of at least two shared hashes from mash screen) were selected and reads were pseudoaligned to this subset of reference genomes using kallisto^60^ (v0.44.0). Kallisto provides two outputs: an “abundances” table containing the number of reads aligned to each gene from the pre-selected set of reference genomes and a pseudo-alignment file (in *.bam format) containing the mapping information for each of the reads processed by kallisto. From the abundances table, we derived a taxonomy table, assigning each gene to its corresponding species, as well as a functional table, mapping each gene to KEGG functional annotation using KEGG Orthology numbers (KOs). To remove potential artifacts, two additional filters were applied to the taxonomic table: first, if less than 10 different functions (i.e. 10 different KOs) were expressed from a given species, such species was discarded. This filter ensures identification of active bacteria, minimizing the capture of contaminants appearing during the sample preparation or sequencing. Second, if one function accounted for more than 95% of the mapped reads of a given species, it was also discarded. This filter was aimed at removing potential artifacts caused by errors in the reference genome assemblies from our database.

Bacterial reads were assigned their specific barcodes and UMIs as follows: read IDs from the mapped microbial reads were retrieved from the kallisto pseudoalignment (*.bam) output using SAMtools (v1.9)^61^. These unique read IDs were used to retrieve the specific barcodes and UMIs using the raw read 1 fastq files, thus assigning each barcode and UMI univocally to a microbial species and function. Barcodes assigned to bacterial species that had been removed in the last two filtering steps of the single-cell analysis pipeline (see above) were discarded, to avoid including potential contaminants in the host-bacteria association analyses.

Differences in lower respiratory tract microbial taxa between COVID-19 patients and controls, ICU and ward patients, and invasive and non-invasive ventilation types were calculated using Wilcoxon rank-sum tests on centered-log-ratio (CLR)-transformed data. This more lenient approach than the one used for 16S data was chosen due to the low number of samples available and the reduced number of bacterial reads identified per sample. Prior to CLR data transformation, we filtered the data using the CoDaSeq.filter function, to keep samples with more than 1,000 reads and taxa with a relative abundance above 0.1%. Zeros were imputed using the same approach as for the 16S amplicon data.

### Direct associations between bacteria and host cells

Host single-cell transcriptomics data was obtained from the Seurat^62^ object after preprocessing and integrating the samples of the single-cell cohort, as described previously^18^. From the Seurat object, the metadata was extracted, including the information on patient group (COVID-19 or control) and severity of the disease (moderate or critical) as well as cell type and subtype annotation corresponding to each barcode. Enrichment of bacteria detected in patient groups or cell types was calculated using chi-squared tests, with effect sizes determined via the standardized residuals. Significance was assessed via post-hoc tests using the R package chisq.posthoc.test^63^.

To evaluate the overlap between bacterial and viral reads detection in host cells of COVID-19 patients, we considered the total number of cells analyzed in these patients: 33,243. Of these, 31,868 cells do not have associated bacterial or viral reads; 1,032 have only bacterial reads; 342 have only viral reads; and 1 has both viral and bacterial reads detected (Supplementary Table 5). The marginal probability for bacterial detection is thus P(bacterial detection) = 1,033/33,243 = 0.031; while the marginal probability for viral detection in this dataset is P(viral detection) = 343/33,243 = 0.010. Assuming independence of both events, the joint probability of finding a host cell associated to both bacterial and viral reads would be P(bacterial and viral detection) = 0.031*0.010 = 3.2·10^−4^. With this joint probability and a total of 33,243 cells profiled, an average of 10.65 host cells should have both bacterial and viral reads detected. A Chi-squared test suggests non-independence of the data (p-value = 4.1·10^−3^). Additionally, we performed an exact binomial test, considering number of successes = 1 (joint bacterial and viral detection), probability of success = 3.2-10^−4^, (the joint probability assuming independence of both events), and number of trials = 33,243 (the total number of cells studied). This two-sided test evaluates the null hypothesis that the joint probability of both events is the one calculated assuming independence. The result of this test (p-value = 5.7·10^−4^) suggests rejecting the null hypothesis. Therefore, these analyses altogether suggest that both events are not independent and that there is mutual exclusion of microbiome members and viruses in the same host immune cells.

For cell types showing an enrichment in associated bacteria, a new Seurat object was created by subsetting the specific cell type. Chi-squared tests were also used to determine enrichment of bacteria-associated cell subtypes. Previous annotations of cell subtypes^18^ were used to generate new clusters manually and identify marker genes for these subtypes, using the function findAllMarkers from Seurat. This function was also used to find differentially expressed genes between bacteria-associated and not-bacteria-associated host cells of each subtype. When using this function, reported adjusted p-values are calculated using Bonferroni correction by default.

## Data Availability

Raw amplicon sequencing data that support the findings of this study have been deposited at the European Genome-phenome Archive (EGA), with accession no EGAS00001004951. The single cell RNA-seq data was first described in a separate publication and deposited also in EGA with accession number EGAS00001004717.

## Supplementary Figure Legends

**Supplementary Figure 1**. Alpha diversity in the upper respiratory tract. a) Shannon diversity index of all samples, stratified by the sampling moment: admission, throughout ICU stay or at ICU discharge/during treatment in ward. The p-value of a Kruskal-Wallis test, as well as the those of Dunn tests corresponding to the pairwise differences among the three groups, are shown. b) Forest plot of the fixed effects estimates of the variables selected in the best model predicting Shannon diversity index. The points and values above indicate the fixed effect estimates of the variables selected, while the horizontal lines span their 95% confidence intervals. Asterisks denote significance as follows: * = p-value ≤ 0.05; ** = p-value ≤ 0.01; *** = p-value ≤ 0.001; **** = p-value ≤ 0.0001. c) c) Effect of the length of ICU stay and SARS-CoV-2 viral load on upper respiratory tract microbiome diversity. Each plot shows the model-predicted Shannon index as a function of the days in ICU, for a different level of SARS-CoV-2 viral load (selected within the range of observed data). Shaded areas correspond to the 95% confidence intervals. d) Association of the length of ICU stay and calprotectin gene expression levels with upper respiratory tract microbiome diversity. Each plot shows the model-predicted Shannon index as a function of the days in ICU, for a different level of calprotectin (subunit S100A8) gene expression, selected within the range of observed data. Shaded areas correspond to the 95% confidence intervals. e) Model-averaged relative importance of each of the variables selected (only for fixed effects). Variable importance was calculated as a weighted average of the models in which each of the variables appeared, with weights corresponding to the model ranks, defined by their AICc values. f) Intra-patient differences of alpha-diversity values, before and after administration of meropenem/piperacillin-tazobactam (left) or mechanical ventilation (right). P-values shown are derived from Wilcoxon signed-rank tests. For (a,f), boxplots span from the first until the third quartile of the data distribution, and the horizontal line indicates the median value of the data. The whiskers extend from the quartiles until the last data point within 1.5 times the interquartile range, with outliers beyond. Individual data points are also represented.

**Supplementary Figure 2**. Association of antibiotics and mechanical ventilation. a) Mosaic plots showing, for each category of oxygen support, the proportion of samples receiving ceftriaxone administration (current administration on day of sampling, left) or the proportion of samples having received meropenem or piperacillin-tazobactam (ongoing or previous treatment, right). P-values denote the significance of Chi-squared tests for these associations. The different oxygen support levels are: 1-oxygen flow (via nasal cannula); 2-high flow oxygen support; 3-non-invasive ventilation (CPAP, BIPAP); 4-invasive ventilation; 5-prone ventilation; 6-extra corporeal membrane oxygenation (ECMO); 7-nitric oxide inhalation. Levels 4-7 correspond to mechanically ventilated patients. b) Longitudinal sampling of patients, showing specific antibiotic administration. Each line represents one patient. Yellow lines represent no-specific antibiotic administration for the spanned period; blue lines represent antibiotic was administered during that period. Shaded areas in light gray represent ICU stay, whilst areas in dark gray represent periods with the patient receiving mechanical ventilation. Crosses indicate the timepoints where swab samples were obtained for microbiome analyses. Individual yellow points at later times represent follow-up visits.

**Supplementary Figure 3**. Differentially abundant taxa between oxygen support types. a) The 29 taxa whose abundance is significantly different between non-invasive and invasive ventilation are represented. b) The 20 taxa whose abundance is significantly different between ventilation types, after controlling for antibiotic usage, are represented. Boxplots span from the first until the third quartile of the data distribution, and the horizontal line indicates the median value of the data. The whiskers extend from the quartiles until the last data point within 1.5 times the interquartile range, with outliers beyond. Individual data points are also represented. Gray lines join samples pertaining to the same patient, taken at different time points.

**Supplementary Figure 4**. Absolute microbial read counts in single-cell RNA-seq data from BAL samples. The top 15 species detected in our analyses are depicted. Samples are grouped by disease type (control for non-COVID-19 pneumonia patients, or COVID-19) and hospital stay (ICU or ward).

**Supplementary Figure 5**. Associations of specific cell types with bacteria, for COVID-19 and control samples. The colors represent the strength of the association as the standardized residuals of a Chi-squared test. Red colors indicate a positive association (i.e. enrichment) of bacteria for each cell type. Blue colors indicate a negative association (i.e. depletion) of bacteria for a given cell type. Asterisks denote significance as follows: * = p-value ≤ 0.05; ** = p-value ≤ 0.01; *** = p-value ≤ 0.001; **** = p-value ≤ 0.0001.

## Author contributions

VLR, ACG, JW, JR designed the study. SJ, TVW, JN, CD, JG, GH, PM collected and processed the BAL samples. PVM and LV collected the clinical data. JW and EW collected the swabs. VLR, ACG and JVW processed the swabs. JVW, MB and SMM determined SARS-CoV-2 viral loads and host gene expression from swabs. DL and JQ generated the single-cell raw data as well as the processed gene-count matrix with annotations of cell types and subtypes. VLR and AGC analyzed the data. VLR, ACG and JR wrote the manuscript. All authors have read and approved the manuscript.

## Acknowledgments

This study has been supported by funding from the VIB Grand Challenges Program. VLR is supported by an FWO senior postdoctoral fellowship (12V9421N). ACG is supported by an EMBO postdoctoral fellowship (ALTF 349-2019). The Raes lab is supported by KU Leuven, the Rega institute and VIB.

## CONTAGIOUS collaborators

Yannick Van Herck, Alexander Wilmer, Michael Casaer, Stephen Rex, Nathalie Lorent, Jona Yserbyt, Dries Testelmans, Karin Thevissen.

## Data availability statement

Raw amplicon sequencing data that support the findings of this study have been deposited at the European Genome-phenome Archive (EGA), with accession no EGAS00001004951. The single cell RNA-seq data was first described in a separate publication^18^ and deposited also in EGA with accession number EGAS00001004717.

